# PTSD-related differences in resting-state functional connectivity and associations with sex hormones

**DOI:** 10.1101/2024.09.26.24314301

**Authors:** Natalie C. Noble, Mohammad S. E. Sendi, Julia B. Merker, Samantha R. Linton, Theresa K. Webber, Amit Etkin, Wei Wu, Kerry J. Ressler, Antonia V. Seligowski

## Abstract

**Background:** Posttraumatic stress disorder (PTSD) is a debilitating condition that disproportionately impacts individuals who are female. Prior research indicates that males with PTSD exhibit hypoconnectivity of frontal brain regions measured with resting electroencephalography (EEG). The present study examined functional connectivity among females with PTSD and trauma-exposed controls, as well as the impact of sex hormones.

**Methods:** Participants included 61 females (*M*age = 31.41, *SD* = 8.64) who endorsed Criterion A trauma exposure. Resting state EEG data were recorded for five minutes in the eyes open position. Using a Linear Mixed Effects model, paired region-of-interest power envelope connectivity of the theta band (4-7 Hz) served as the response variables.

**Results:** Compared to controls, the PTSD group displayed hyperconnectivity between visual brain regions and the rest of the cerebral cortex (*pFDR* < 0.05). Additionally, participants with PTSD demonstrated enhanced connectivity between the default mode network and frontoparietal control network compared to controls (*pFDR* < 0.05), as well as increased connectivity between the ventral attention network and the rest of the cerebral cortex (*pFDR* < 0.05). Estradiol was associated with higher connectivity, while progesterone was associated with lower connectivity, but these did not survive correction.

**Conclusions:** Results are consistent with prior research indicating that PTSD is associated with altered connectivity in visual brain regions, which may reflect disrupted visual processing related to reexperiencing symptoms (e.g., intrusive memories). Our findings provide additional support for the relevance of the theta frequency range in PTSD given its role in fear learning and regulation processes.

## Introduction

Posttraumatic stress disorder (PTSD) is a debilitating condition that involves re-experiencing symptoms, avoidance of trauma reminders, negative changes in thinking and mood, and altered arousal following a traumatic event (American Psychiatric Association, 2013). In the U.S., PTSD affects roughly 12 million individuals in a given year, equating to an annual healthcare cost of approximately $230 billion (Davis et al., 2022). Both magnetic resonance imaging (MRI) and electroencephalography (EEG) techniques have captured important insights into altered fear processing among individuals with PTSD. Despite these advancements, little research has been conducted in female samples, a population that faces a twofold risk of developing PTSD (Kilpatrick et al., 2013).

Numerous MRI-based studies have demonstrated that PTSD is associated with hypoactivity of frontal brain regions, such as the ventromedial prefrontal cortex (vmPFC; Falconer et al., 2008; Milad et al., 2009; Rougemont-Bücking et al., 2011). Additionally, MRI-based studies have captured reduced connectivity between the vmPFC and limbic structures involved in the fear response, such as the amygdala and hippocampus (Spirada et al., 2012; Jin et al., 2013). As reviewed by Koch and colleagues (2016), hypoactivation of the vmPFC among individuals with PTSD is thought to reflect diminished top-down regulation, or inhibition, over the fear response during non-threatening situations. Consistent with clinical presentations of PTSD, unchecked activation of the amygdala is associated with hypervigilance and hyperarousal (Rauch et al., 2006).

Several high-density EEG studies have now replicated MRI-based studies in PTSD. For example, a resting-state EEG study of civilians with PTSD demonstrated decreased functional connectivity of frontal brain regions within the beta and gamma frequency bands (Lee et al., 2014). Similarly, in their resting-state EEG study of male combat Veterans with PTSD, Toll and colleagues (2020) observed hypoconnectivity of the orbital and anterior middle frontal gyri, structures located in the frontal lobe. These findings were detected with a novel analytical technique involving source-localization and orthogonalization of amplitude correlations. Importantly, the results were found to be significant in the theta frequency band (4-7 Hz), which has been implicated in fear learning (Mueller et al., 2014; Sperl et al., 2018).

In females, PTSD has been associated with decreased vmPFC activity (Jovanovic, 2013), disrupted vmPFC-amygdala connectivity (Stevens et al., 2013), and decreased connectivity between the posterior cingulate cortex and the precuneus, vmPFC, hippocampus, and amygdala (Bluhm et el., 2009). With the exception of the amygdala, these structures are thought to contribute to the default mode network, which is typically activated during introspective tasks, such as self-reflection (Aguilar & McNally, 2022; Philippi & Koenigs, 2014). Estradiol and progesterone have also been associated with neural connectivity, as well as neural markers of stress reactivity (e.g., Hidalgo-Lopez et al., 2020, 2021; Mulligan et al., 2019; Syan et al., 2017; Zhang et al., 2013). For example, Brierworth and colleagues (2021) observed attenuated theta oscillations in the dorsal anterior cingulate cortex among females with high estradiol levels during fear recall compared to those with low estradiol and males. Further research involving trauma-exposed samples is necessary to understand the complex interaction between PTSD and sex hormones.

The current study examined resting functional connectivity among trauma-exposed females with and without PTSD. Based on prior research in male samples, we hypothesized that the PTSD group would demonstrate lower functional connectivity of frontal brain regions compared to controls, and that this difference would emerge for the theta frequency range (4-7 Hz). Given prior research demonstrating that low estradiol levels are associated with worse PTSD severity and greater activation of the dorsal anterior cingulate cortex, we also hypothesized that lower estradiol levels would be associated with lower functional connectivity. We did not have a-priori hypotheses about progesterone given limited prior research in trauma and PTSD samples.

## Methods

Participants included 66 females (*M*age = 31.45, *SD* = 8.92) who endorsed DSM-5 Criterion A trauma exposure. In terms of race, 53 participants identified as White (80.3%), four (6.1%) as Asian or South-Asian, and five (7.6%) as Black or African American; four participants (6.1%) indicated that their race was not listed or chose not to respond. Eight participants (12.3%) identified as being of Latino, Hispanic, or Spanish origin.

Following consent, participants completed psychological measures and provided a blood sample, from which estradiol (pg/mL) and progesterone (ng/mL) using were assayed using mass spectrometry. Participants were then prepared for the EEG measurement. The Institutional Review Board approved all study procedures and participants received $100 as compensation.

### Psychological measures

A demographics questionnaire was used to assess age, race/ethnicity, gender identity, marital status, education, employment, and income. Trauma exposure was measured with the Life Events Checklist (Weathers et al., 2013), which is a self-report measure of 17 types of potentially traumatic events (e.g., natural disaster, sexual assault). PTSD was assessed with the Clinician-Administered PTSD Scale for DSM-5 (CAPS-5; Weathers et al., 2012).

### EEG data acquisition

Resting state data were recorded for five minutes in the eyes open position using a 128-channel Electrical Geodesics saline EEG system. Participants were asked to remain still in order to minimize eye blinks and movements, and they were asked to look at a fixation cross. Electrode placement was in the standard 10-20 system. Data were collected at 1000 Hz with 0.1-100 Hz analog filtering, using Cz as a reference. Impedances were kept below 100 kΩ.

### EEG data preprocessing

EEG data were preprocessed using a fully automated artifact rejection pipeline that has been validated in our prior work (Toll et al., 2020) and minimizes bias from subjective manual artifact rejection. The preprocessing pipeline involved: 1) resampling data to 250 Hz, 2) removal of 60 Hz a.c. line noise artifact, 3) removal of non-physiological low frequency data using a 0.01 Hz high-pass filter, 4) rejection of bad epochs by thresholding the magnitude of each epoch, 5) rejection of bad channels by thresholding spatial correlations among channels, 6) exclusion of participants with more than 25% bad channels, 7) estimation of EEG signals from bad channels from the adjacent channels through spherical spline interpolation, 8) independent components analysis to remove remaining artifacts, including scalp muscle artifact, ocular artifact, and electrocardiogram artifact, 9) re-referencing signals to the common average, and 10) filtering of signals to four frequency ranges: theta (4–7 Hz), alpha (8–12 Hz), beta (13–30 Hz), and low gamma (31–50 Hz).

### EEG connectivity processing

Complex-valued time series data were analyzed from each vertex in the brain’s source-space. Each vertex’s analytical signal was orthogonally separated from every other vertex, a process that helps in isolating signals from each brain region. We then calculated the power envelopes from these orthogonalized time series and used the natural logarithm to normalize them. We then computed Pearson’s correlation coefficients between the log-transformed power envelopes of each pair of vertices. This method was compared to another where we analyzed connectivity matrices based on raw power envelopes from non-orthogonalized time series. We focused our connectivity analysis on 31 regions of interest within the Montreal Neurological Institute space. These regions included the left and right Visual Area 1 (V1), Somatosensory Cortex (SMC), Inferior Frontal Junction (IFJ), Intraparietal Sulcus (IPS), Frontal Eye Fields (FEF), Supplemental Eye Fields (SEF), Angular Gyrus (ANG), Posterior Middle Frontal Gyrus (PMFG), Inferior Parietal Lobule (IPL), Orbital Gyrus (ORB), Middle Temporal Gyrus (MTG), Anterior Middle Frontal Gyrus (AMFG), Insula (INS), and Supramarginal Gyrus (SUP), as well as the Posterior Cingulate Cortex (PCC), Medial Prefrontal Cortex (mPFC), Dorsal Anterior Cingulate Cortex (DACC). These regions were selected based on an independent parcellation from a previous study’s analysis (Chen et al., 2013). This approach allowed us to ground our EEG regions of interest in a well-established, fMRI-based framework for major cortical connectivity networks, while adjusting for the lower resolution of EEG data. We then applied the Fisher z transformation to these correlations, a statistical method that normalizes the data for better comparison and analysis.

### Data analysis

For each brain connectivity measurement, we identified the central data range using the 25^th^ and 75th percentiles. Then, we calculated the interquartile range and established outlier thresholds at 1.5 times the interquartile range above and below these percentiles. Data points outside these boundaries were marked as outliers and excluded from further analysis. Next, using a linear mixed effects model, paired region-of-interest power envelope connectivity of the theta band (4- 7 Hz) served as the response variables. The model included PTSD diagnosis as a categorical predictor, and age, marital status, education, and income as covariates. A random intercept for each participant was also incorporated into the model. P-values derived from this analysis were corrected using a single false discovery rate adjustment across all regions of interest using the Benjamini-Hochberg method (pFDR, 465 comparisons < 0.05).

To explore the relationship between theta band power envelope connectivity and levels of estradiol and progesterone, we employed partial correlations. This analysis covaried for age, marital status, education level, and income. In all statistical evaluations, p-values were adjusted using the Benjamini-Hochberg correction method to account for multiple comparisons. This adjustment was made across 465 comparisons, with a significance threshold set at pFDR < 0.05.

## Results

Thirty-seven (56.9%) participants met diagnostic criteria for PTSD per the CAPS-5. Compared to controls, participants with PTSD demonstrated significantly increased connectivity (pFDR < 0.05) between visual brain regions and other areas of the cerebral cortex (**Figure 1**). Specifically, there was a notable increase in connectivity from the left visual brain regions to various key areas: the right supplementary eye fields, part of the dorsal attention network; the left angular gyrus, associated with the default mode network; the left posterior middle frontal gyrus, left inferior parietal lobule, and left middle temporal gyrus, all of which are components of the frontoparietal control network; and the left supramarginal gyrus, involved in the ventral attention network. Similarly, enhanced connectivity was observed between the right visual region and left angular gyrus, left inferior parietal lobule, and left supramarginal gyrus, in the PTSD group compared to the control group.

**Figure 1:**
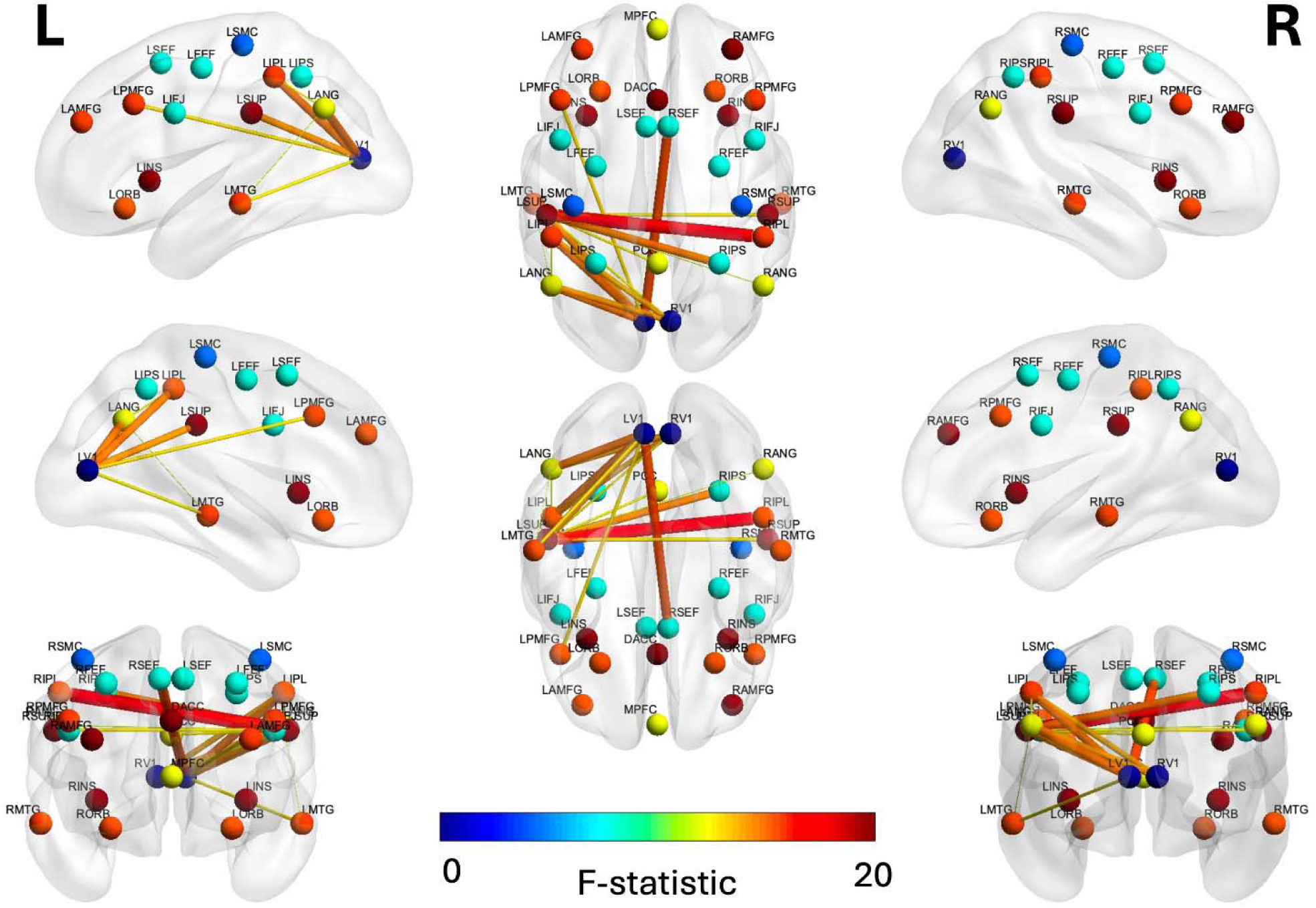
**Theta band hyperconnectivity in PTSD.** *Note.* Thirty-one regions of interest were defined in Montreal Neurological Institute space. We created linear mixed-effects models for the functional connectivity between 465 unique pairs of the 31 regions and diagnosis (i.e., trauma-exposed versus PTSD) in the theta band, incorporating age, marital status, education, and income as covariates. A random intercept for each participant was also incorporated into the model. The F-statistics of the models associated with each connectivity pair that survived after false discovery rate correction are shown from different views. The thickness and color of the edges in the figure represent the strength of the F-value. The node colors represent the network to which the node belongs. **LV1**: Left Visual Area 1 (Primary Visual Cortex); **RV1**: Right Visual Area 1 (Primary Visual Cortex); **LSMC**: Left Somatosensory Cortex; **RSMC**: Right Somatosensory Cortex; **LIFJ**: Left Inferior Frontal Junction; **RIFJ**: Right Inferior Frontal Junction; **LIPS**: Left Intraparietal Sulcus; **RIPS**: Right Intraparietal Sulcus; **LFEF**: Left Frontal Eye Field; **RFEF**: Right Frontal Eye Field; **LSEF**: Left Supplementary Eye Field; **RSEF**: Right Supplementary Eye Field; **PCC**: Posterior Cingulate Cortex; **MPFC**: Medial Prefrontal Cortex; **LANG**: Left Angular Gyrus; **RANG**: Right Angular Gyrus; **LPMFG**: Left Posterior Middle Frontal Gyrus; **RPMFG**: Right Posterior Middle Frontal Gyrus; **LIPL**: Left Inferior Parietal Lobule; **RIPL**: Right Inferior Parietal Lobule; **LORB**: Left Orbital Gyrus; **RORB**: Right Orbital Gyrus; **LMTG**: Left Middle Temporal Gyrus; **RMTG**: Right Middle Temporal Gyrus; **LAMFG**: Left Anterior Middle Frontal Gyrus; **RAMFG**: Right Anterior Middle Frontal Gyrus; **LINS**: Left Insula; **RINS**: Right Insula; **DACC**: Dorsal Anterior Cingulate Cortex; **LSUP**: Left Supramarginal Gyrus; **RSUP**: Right Supramarginal Gyrus.

In addition, we noted increased connectivity in the PTSD group compared to the control group between several additional brain regions. This includes heightened connectivity from the left supramarginal gyrus, which is a component of the ventral attention network, to various areas: the right intraparietal sulcus, part of the dorsal attention network; the posterior cingulate cortex and the right angular gyrus, both associated with the default mode network; the right inferior parietal lobule, a component of the frontoparietal control network; and the right supramarginal gyrus, also part of the ventral attention network.

As shown in **Figure 2**, higher estradiol levels were associated with higher theta band connectivity in multiple brain regions, particularly those in the default mode network. The most substantial correlation was observed between the mPFC and the left intraparietal sulcus. Specifically, higher estradiol levels were associated with stronger connectivity between the mPFC and the left angular gyrus, *r* = .42 (uncorrected *p* = .003). In addition, higher estradiol levels were associated with stronger connectivity between the mPFC and the left angular gyrus, *r* = .41 (uncorrected *p* = .003). No significant associations were identified following FDR correction.

**Figure 2:**
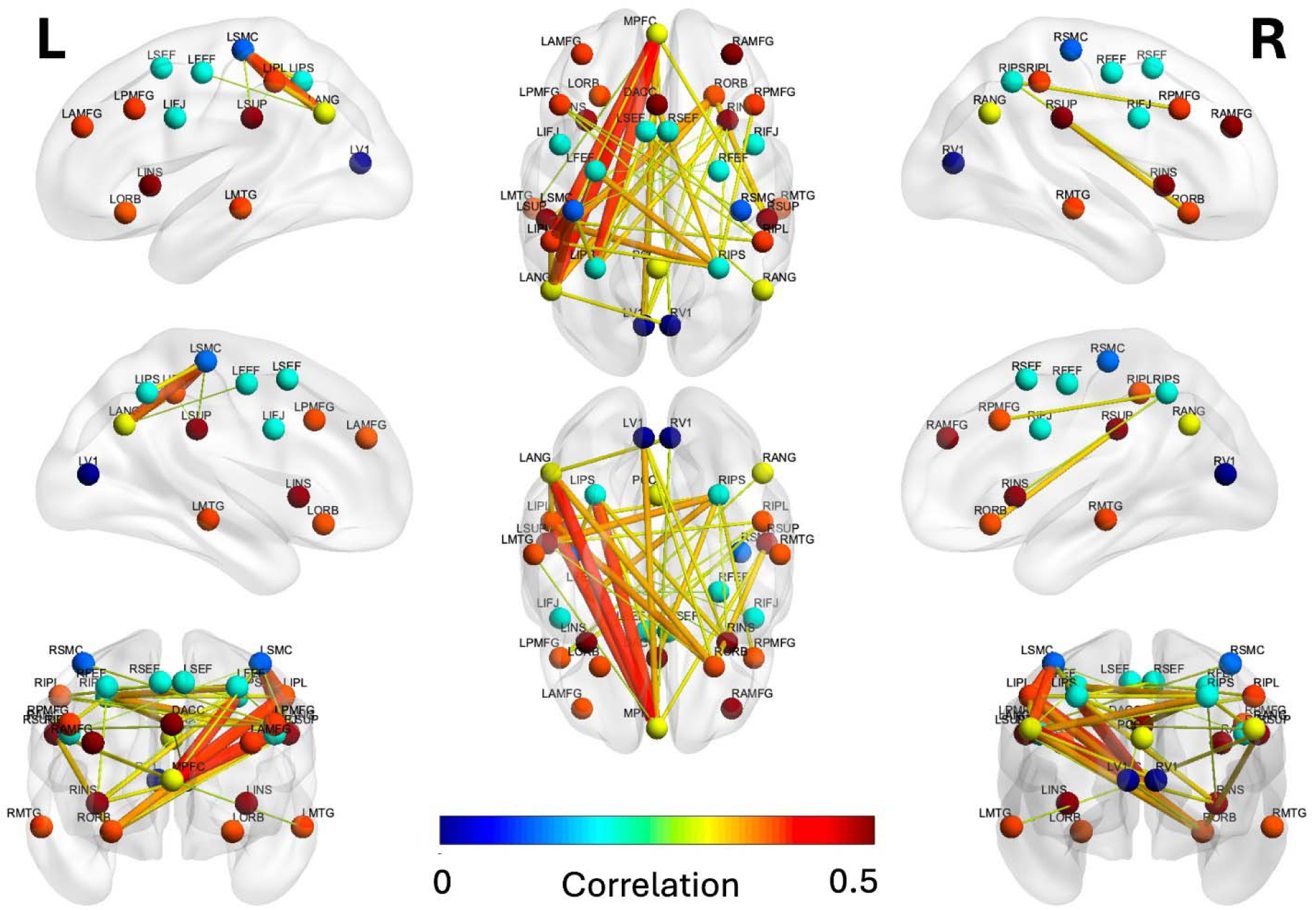
**Higher estradiol levels associated with higher theta band connectivity.** *Note.* A partial correlation was performed between theta band connectivity among 31 regions of interest, resulting in 465 unique connections, and estradiol levels, accounting for age, marital status, education, and income as covariates. Only correlation values with associated p-values less than 0.05 are shown. The thickness and color of the edges represent the strength of the F-value, while the node colors indicate the network to which the node belongs. **LV1**: Left Visual Area 1 (Primary Visual Cortex); **RV1**: Right Visual Area 1 (Primary Visual Cortex); **LSMC**: Left Somatosensory Cortex; **RSMC**: Right Somatosensory Cortex; **LIFJ**: Left Inferior Frontal Junction; **RIFJ**: Right Inferior Frontal Junction; **LIPS**: Left Intraparietal Sulcus; **RIPS**: Right Intraparietal Sulcus; **LFEF**: Left Frontal Eye Field; **RFEF**: Right Frontal Eye Field; **LSEF**: Left Supplementary Eye Field; **RSEF**: Right Supplementary Eye Field; **PCC**: Posterior Cingulate Cortex; **MPFC**: Medial Prefrontal Cortex; **LANG**: Left Angular Gyrus; **RANG**: Right Angular Gyrus; **LPMFG**: Left Posterior Middle Frontal Gyrus; **RPMFG**: Right Posterior Middle Frontal Gyrus; **LIPL**: Left Inferior Parietal Lobule; **RIPL**: Right Inferior Parietal Lobule; **LORB**: Left Orbital Gyrus; **RORB**: Right Orbital Gyrus; **LMTG**: Left Middle Temporal Gyrus; **RMTG**: Right Middle Temporal Gyrus; **LAMFG**: Left Anterior Middle Frontal Gyrus; **RAMFG**: Right Anterior Middle Frontal Gyrus; **LINS**: Left Insula; **RINS**: Right Insula; **DACC**: Dorsal Anterior Cingulate Cortex; **LSUP**: Left Supramarginal Gyrus; **RSUP**: Right Supramarginal Gyrus.

As shown in **Figure 3**, lower progesterone levels were associated with higher theta band connectivity in multiple brain regions. Specifically, lower progesterone levels were associated with stronger connectivity between the right somatosensory cortex and the right frontal eye fields, *r* = -.35 (uncorrected *p* = .024). In addition, lower progesterone levels were associated with stronger connectivity between the right somatosensory cortex and the right anterior middle frontal gyrus, *r* = -.31 (uncorrected *p* = .043), and between the right supplementary eye fields and the right inferior parietal lobule, *r* = -.30 (uncorrected *p* = .0497). No significant associations were identified following FDR correction.

**Figure 3:**
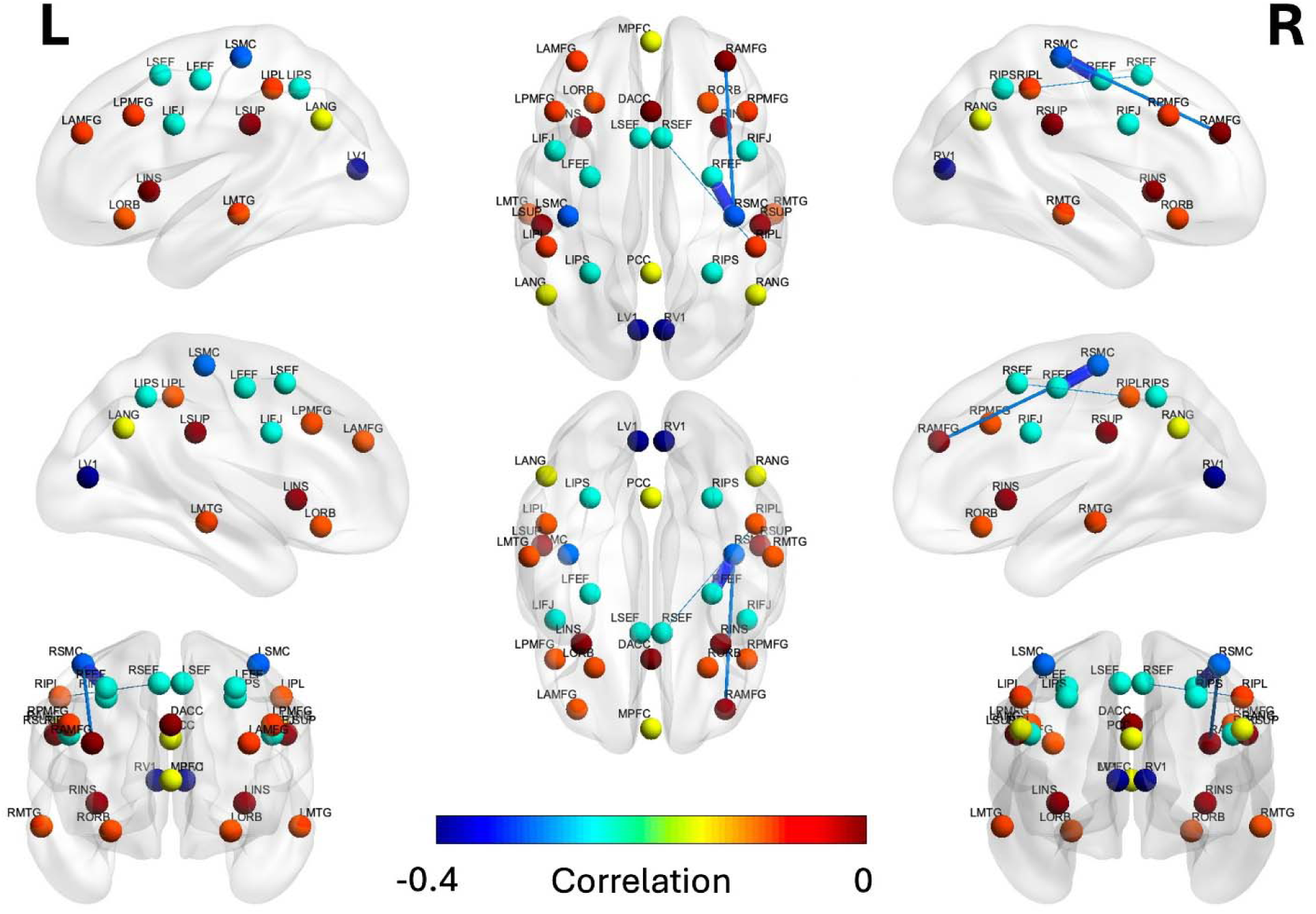
**Lower progesterone levels associated with higher theta band connectivity.** *Note.* A partial correlation was performed between theta band connectivity among 31 regions of interest, resulting in 465 unique connections, and progesterone levels, accounting for age, marital status, education, and income as covariates. Only correlation values with associated p-values less than 0.05 are shown. The thickness and color of the edges represent the strength of the F-value, while the node color indicates the network to which the node belongs. **LV1**: Left Visual Area 1 (Primary Visual Cortex); **RV1**: Right Visual Area 1 (Primary Visual Cortex); **LSMC**: Left Somatosensory Cortex; **RSMC**: Right Somatosensory Cortex; **LIFJ**: Left Inferior Frontal Junction; **RIFJ**: Right Inferior Frontal Junction; **LIPS**: Left Intraparietal Sulcus; **RIPS**: Right Intraparietal Sulcus; **LFEF**: Left Frontal Eye Field; **RFEF**: Right Frontal Eye Field; **LSEF**: Left Supplementary Eye Field; **RSEF**: Right Supplementary Eye Field; **PCC**: Posterior Cingulate Cortex; **MPFC**: Medial Prefrontal Cortex; **LANG**: Left Angular Gyrus; **RANG**: Right Angular Gyrus; **LPMFG**: Left Posterior Middle Frontal Gyrus; **RPMFG**: Right Posterior Middle Frontal Gyrus; **LIPL**: Left Inferior Parietal Lobule; **RIPL**: Right Inferior Parietal Lobule; **LORB**: Left Orbital Gyrus; **RORB**: Right Orbital Gyrus; **LMTG**: Left Middle Temporal Gyrus; **RMTG**: Right Middle Temporal Gyrus; **LAMFG**: Left Anterior Middle Frontal Gyrus; **RAMFG**: Right Anterior Middle Frontal Gyrus; **LINS**: Left Insula; **RINS**: Right Insula; **DACC**: Dorsal Anterior Cingulate Cortex; **LSUP**: Left Supramarginal Gyrus; **RSUP**: Right Supramarginal Gyrus.

## Discussion

Our findings provide additional support for the relevance of the theta frequency range in PTSD given its role in fear learning and regulation processes (Mueller et al., 2014; Sperl et al., 2018). We observed hyperconnectivity of frontal brain regions among our female sample, which is in contrast to prior research in male Veterans (Toll et al., 2020), and we replicated prior work demonstrating altered visual connectivity among trauma-exposed populations (Harnett et al., 2022a, 2022b; Rowland et al., 2023). Further, our results are consistent with prior research indicating that PTSD is associated with altered connectivity within and between the default mode network, ventral attention network, and frontoparietal control network (Lanius et al., 2010). Our results also suggest that estradiol and progesterone may have different associations with neural activity in trauma-exposed females, regardless of PTSD status.

Consistent with prior literature, the present findings indicated altered connectivity in visual brain regions for individuals with PTSD. One prior study of predominantly male Veterans reported a relationship between greater re-experiencing symptoms and both increased visual-sensorimotor and decreased visual-frontoparietal functional connectivity (Maron-Katz et al., 2020). In our study, the right and left visual brain regions demonstrated increased connectivity with regions of the default mode network, frontoparietal control network, and ventral attention network, as well as with regions of the dorsal attention network for the left visual brain region only. These alterations may reflect the aforementioned disrupted visual processing related to PTSD symptoms, and re-experiencing in particular, which has recently been shown in acutely trauma- exposed individuals (Harnett et al., 2022a, 2022b; Rowland et al., 2023). Further investigation is required to determine the precise nature of this relationship among females with PTSD.

Our results indicated greater connectivity between several brain regions for females with PTSD. Specifically, we observed increased connectivity *within* the ventral attention (i.e., salience) network as well as between the ventral attention network and various regions of the dorsal attention network, default mode network, and frontoparietal control (i.e., central executive) network. These findings are consistent with a prior study that observed a relationship between salience network connectivity and hyperarousal symptoms in PTSD, and it has been posited that those with PTSD are overly “primed” to detect salience (e.g., threat) in their environment (Akiki et al., 2017). Although hyperconnectivity between the ventral attention and frontoparietal control networks has not previously been reported in PTSD, one prior study observed hyperconnectivity between these networks within the context of a subliminal threat-processing paradigm (Rabellino et al., 2015). Finally, our observation of *hyper*connectivity of frontal brain regions is in contrast to a study in male Veterans that observed *hypo*connectivity of these regions (Toll et al., 2020), which emphasizes the need for sex-based comparisons that may shed light on sex-specific neural alterations in PTSD.

These results illustrate altered connectivity across several key regions of interest within the theta range, which has been implicated in fear learning and regulation (Mueller et al., 2014; Sperl et al., 2018). Given the high prevalence of altered fear learning and general cognitive dysfunction in PTSD (Dunkley et al., 2015), investigations of connectivity within the theta range may offer unique insights into the neurobiological underpinnings of this relationship. It is also important to note that alpha-band oscillations have been identified as the dominant (i.e., easiest to detect) oscillations in the human brain during resting wakefulness (Klimesch, 2012). In the present study, alterations in connectivity during resting wakefulness were tested and observed only within the theta range. This suggests that the alterations in connectivity cannot purely be accounted for by the ease with which the signal was detected, which supports the conclusion that this altered connectivity reflects true differences between the PTSD and trauma-exposed control groups.

In terms of sex hormones, higher estradiol levels were associated with higher connectivity between the mPFC and the left intraparietal sulcus – in the default mode network. While no prior research tested this in trauma-exposed females, our finding is consistent with a study that demonstrated higher neural connectivity in females during the mid-luteal phase of the menstrual cycle (when estradiol levels are higher; Hidalgo-Lopez et al., 2020). Given that greater connectivity in the default mode network is associated with decreased PTSD severity, this finding is also consistent with literature suggesting a protective effect of estradiol (Akiki et al., 2018). In contrast, lower progesterone levels were associated with higher connectivity between the right somatosensory cortex and regions of both the dorsal attention and salience networks, as well as between regions of the dorsal attention and frontoparietal control networks. While this relationship has not previously been explored in trauma-exposed females, a prior study of healthy women found greater neural connectivity within the salience network for individuals with lower relative progesterone (Hidalgo-Lopez et al., 2021). Of note, the hormonal associations identified in the present study did not survive correction for multiple testing and require replication in larger samples.

A strength of this study was our use of high-density EEG, coupled with a novel analytical technique involving source-localization and orthogonalization, which enabled us to examine the functional connectivity of 465 ROI pairs with enhanced spatial resolution. Second, our trauma- exposed female sample allowed us to explore potential neural deficits related to PTSD among a high-risk, underrepresented demographic. Further, our study utilized the CAPS-5, which is considered the gold-standard in PTSD assessment. In terms of limitations, our lack of a male sample precluded us from making any sex-based comparisons of neural connectivity. In addition, our interpretation is complicated by the inclusion of hormonal data from non-naturally cycling participants (i.e., those taking hormonal birth control). Although biological assays provide an accurate measure of circulating gonadal hormone levels, the implications of exogenous hormone administration on the relationship between gonadal hormone levels and symptoms of psychopathology are not well understood. Future studies should therefore seek to exclusively recruit naturally cycling females.

This is the first study to examine associations among sex hormone levels and neural connectivity in trauma-exposed females. The observed alterations in theta-based connectivity provide additional support for the role of theta in fear learning and regulation. Further, our EEG-based connectivity findings of visual cortex alterations and enhanced visual sensitization in PTSD complement and validate prior MRI findings. Consistent with prior literature, our results also suggest that higher relative estradiol levels may be associated with a protective effect, marked by greater neural connectivity and decreased PTSD symptomatology. Given that this association did not remain significant following correction for multiple comparisons, these findings require replication. Future studies are needed to compare these findings in larger mixed-sex samples.

## Data Availability

All data produced in the present study are available upon reasonable request to the authors

## Funding and Disclosures

AVS supported by K23MH125920 and K23MH125920-03W1; MSES supported by T32MH125786 and receives consulting fees from NIJI Corp. AE reports equity and salary from Alto Neuroscience and equity in Akili Interactive. KJR serves on Scientific Advisory Boards for Sage, Boehringer Ingelheim, Senseye, and the Brain Research Foundation, and has received sponsored research support from Alto Neuroscience. AVS, NCN, JBM, SRL, TKW, and WW report no biomedical financial interests or potential conflicts of interest.

## Notes

### Competing Interest Statement

MSES receives consulting fees from NIJI Corp. AE reports equity and salary from Alto Neuroscience and equity in Akili Interactive. KJR serves on Scientific Advisory Boards for Sage, Boehringer Ingelheim, Senseye, and the Brain Research Foundation, and has received sponsored research support from Alto Neuroscience. AVS, NCN, JBM, SRL, TKW, and WW report no biomedical financial interests or potential conflicts of interest.

### Funding Statement

MSES supported by T32MH125786; AVS supported by K23MH125920 and K23MH125920-03W1.

### Author Declarations

The Mass General Brigham Institutional Review Board approved all study procedures.

